# Whole-exome sequencing in 16,511 individuals reveals a role of the HTRA1 protease and its substrate EGFL8 in brain white matter hyperintensities

**DOI:** 10.1101/2021.03.26.21253954

**Authors:** Rainer Malik, Nathalie Beaufort, Simon Frerich, Benno Gesierich, Marios K Georgakis, Kristiina Rannikmäe, Amy C Ferguson, Christof Haffner, Matthew Traylor, Michael Ehrmann, Cathie LM Sudlow, Martin Dichgans

## Abstract

White matter hyperintensities (WMH) are among the most common radiological abnormalities in the ageing population and an established risk factor for stroke and dementia. While common variant association studies have revealed multiple genetic loci with an influence on WMH volume, the contribution of rare variants to WMH burden in the general population remains largely unexplored. We conducted a comprehensive analysis of WMH burden in the UK Biobank using publicly available whole-exome sequencing data (N=16,511) and found a splice-site variant in *GBE1*, encoding 1,4-alpha-glucan branching enzyme 1, to be associated with lower white matter burden on an exome-wide level (c.691+2T>C, beta=-0.74, se=0.13, p=9.7E-9). Applying whole-exome gene-based burden tests, we found damaging missense and loss-of-function variants in *HTRA1* to associate with increased WMH volume (p=5.5E-6, FDR=0.04). *HTRA1* encodes a secreted serine protease implicated in familial forms of small vessel disease. Domain-specific burden tests revealed that the association with WMH volume was restricted to rare variants in the protease domain (amino acids 204-364; beta=0.79, se=0.14, p=9.4E-8). The frequency of such variants in the UK Biobank population was 1 in 450. WMH volume was brought forward by approximately 11 years in carriers of a rare protease domain variant. A comparison with the effect size of established risk factors for WMH burden revealed that the presence of a rare variant in the HTRA1 protease domain corresponded to a larger effect than meeting the criteria for hypertension (beta=0.26, se=0.02, p=2.9E-59) or being in the upper 99.8% percentile of the distribution of a polygenic risk score based on common genetic variants (beta=0.44, se=0.14, p=0.002). In biochemical experiments, most (6/9) of the identified protease domain variants resulted in a markedly reduced protease activity. We further found *EGFL8*, which showed suggestive evidence for association with WMH volume (p=1.5E-4, FDR=0.22) in gene burden tests, to be a direct substrate of HTRA1 and to be preferentially expressed in cerebral arterioles and arteries. In a phenome-wide association study (PheWAS) mapping ICD-10 diagnoses to 741 standardized Phecodes, rare variants in the HTRA1 protease domain were associated with multiple neurological and non-neurological conditions including migraine with aura (OR=12.24, 95%CI [2.54-35.25], p=8.3E-5). Collectively, these findings highlight an important role of rare genetic variation and of the HTRA1 protease in determining WMH burden in the general population.

## INTRODUCTION

White matter hyperintensities (WMH) are common in elderly individuals (Garde *et al*., 2000; Debette and Markus, 2010) and an increasingly recognized risk factor for stroke (Wardlaw *et al*., 2013; Debette *et al*., 2019), dementia (Wardlaw *et al*., 2013; Dichgans and Leys, 2017; Debette *et al*., 2019), and functional decline in the older age (Ladis Study Group, 2011). They are further associated with poor long-term outcomes after ischemic stroke including an increased risk of stroke recurrence, dementia, and mortality (Georgakis *et al*., 2019). While not specific for any particular etiology, WMH are considered a marker of cerebral small vessel disease (SVD) (Dichgans and Leys, 2017; Wardlaw *et al*., 2019). Hypertension is the single strongest treatable risk factor for WMH (Dufouil *et al*., 2001; Wardlaw *et al*., 2013; Georgakis *et al*., 2019). However, the precise mechanisms underlying WMH remain largely elusive.

WMH are highly heritable with estimates ranging from 18-54% (Duperron *et al*., 2018; Persyn *et al*., 2020). Accordingly, recent genome-wide association studies (GWAS) found common genetic variants at multiple loci to be associated with WMH burden (Fornage *et al*., 2011; Armstrong *et al*., 2020; Persyn *et al*., 2020; Sargurupremraj *et al*., 2020). Besides confirming a causal link with hypertension (Sargurupremraj *et al*., 2020) these studies also pinpoint specific molecular pathways and biological mechanisms. Among the most prominent themes are perturbations of the extracellular matrix (ECM) as evidenced by associations at loci that encode matrisome proteins. These loci include *COL4A2* (encoding collagen type IV alpha 2 chain), *EFEMP1* (encoding EGF containing fibulin extracellular matrix protein 1), *VCAN* (encoding versican), and *NID2* (encoding nidogen). Additional themes, that have emerged from recent GWAS, include dysfunction of vascular endothelial and mural cells, the blood-brain barrier, and inflammatory mechanisms (Neurology Working Group of the Cohorts for Heart Aging Research in Genomic Epidemiology Consortium *et al*., 2016; Persyn *et al*., 2020; Sargurupremraj *et al*., 2020). The results from recent GWAS on WMH further highlight links with small vessel stroke, ischemic stroke, intracerebral hemorrhage, and neurodegenerative disease (Malik *et al*., 2018; Sargurupremraj *et al*., 2020; Traylor *et al*., 2021). As such, large scale genetic studies have been instrumental in uncovering core pathways and mechanisms underlying WMH and defining relationships with related phenotypes, in particular SVD.

A close link between WMH and SVD is further supported by observations in familial forms of SVD (Dichgans *et al*., 2019). WMH are a regular feature in carriers of a pathogenic mutation in *NOTCH3* (encoding neurogenic locus notch homolog protein 3), the gene implicated in cerebral autosomal dominant arteriopathy with subcortical infarcts and leukoencephalopathy (CADASIL) (Joutel *et al*., 1996; Dichgans *et al*., 1998). They are further seen in patients with cerebral autosomal recessive arteriopathy with subcortical infarcts and leukoencephalopathy (CARASIL), a severe type of SVD caused by homozygous or bi-allelic *HTRA1* mutations (Hara *et al*., 2009). *HTRA1* encodes a secreted serine protease that associates with the ECM and has been shown to proteolytically process various molecular constituents of the ECM including latent TGF-ß binding protein, vitronectin, and elastin (Beaufort *et al*., 2014). The outstanding importance of perturbations of the ECM in the pathophysiology of WMH is further illustrated by highly penetrant mutations in *COL4A1* and *COL4A2* (Gould et al., 2006; Verdura et al., 2016; Jeanne and Gould, 2017). These mutations cause a broad range of phenotypes including WMH, small vessel stroke, intracerebral hemorrhage, porencephalopathy, and extracerebral manifestations (Verdura *et al*., 2016; Jeanne and Gould, 2017). Notably, extracerebral manifestations such as ocular and renal manifestations are a common feature in various forms of hereditary SVD (Rannikmae *et al*., 2020).

While the above studies have contributed to the understanding of common genetic variation in WMH and the role of highly penetrant mutations in familial SVD, the significance of rare genetic variation for WMH in the general population remains largely unexplored. To the best of our knowledge, previous studies focused on variants included on HumanExome BeadChip arrays (Jian *et al*., 2018) and on genes implicated in familial SVD (Mishra *et al*., 2019; Rutten *et al*., 2020; Cho *et al*., 2021). Of note, some of them included samples that had been selected on the basis of extreme phenotypes (Mishra *et al*., 2019).

The UK Biobank (UKB) (Bycroft *et al*., 2018) is a large-scale (∼500.000 participants) prospective community-based study that recruited from the general midlife population aged between 40 and 69 years and offers phenotypic information on multiple traits including brain imaging with quantitative data on WMH volumes available in about 44,000 individuals (Littlejohns *et al*., 2020). The UKB further offers detailed clinical information mapped to international classification of diseases (ICD-10) codes, genome-wide genotyping of single nucleotide polymorphisms (SNPs), and as of recently, whole-exome sequencing (WES) data. In contrast to genome-wide genotyping, WES enables comprehensive analysis of all genetic variation in coding regions, including rare variants that have previously been inaccessible. In many complex traits and diseases, studying to this variation has contributed to understanding their genetic basis (Flannick *et al*., 2019; Jurgens *et al*., 2020; Wang *et al*., 2020).

Here, we leveraged data from the UKB to systemically investigate associations between rare variation and WMH burden in a population-based setting. Specifically, we set out to: i) identify single variants associated with WMH burden at an exome-wide level, ii) identify genes associated with WMH load in burden tests, iii) experimentally determine the functional consequences of selected variants; iv) identify genes implicated in pathways and interaction networks relevant to SVD; and v) explore the phenotypic spectrum of alternative allele carriers in a phenome-wide association study (PheWAS).

## MATERIALS AND METHODS

This analysis was carried out under the UKB project 2532.

### UK Biobank population

Our resource was the UK Biobank Exome 200k release from October 2020. Primary and secondary analyses were performed with an updated Functional Equivalence (FE) protocol that retains original quality scores in the CRAM files (referred to as the OQFE protocol) (Szustakowski *et al*., 2020).

The 200k release encompasses 1,135 parent-offspring pairs, 3,855 full-sibling pairs, including 101 trios, 27 monozygotic twin pair and 7,461 second degree genetically determined relationships. To avoid bias due to relatedness, we selected an unrelated set of individuals up to a 2^nd^ degree (KING cut-off 0.0877) (Manichaikul *et al*., 2010). For the WMH volume analysis, we preferentially retained individuals with more extreme trait values (i.e. further away from the mean) in the analysis using PRIMUS (Staples *et al*., 2014). For PheWAS analyses, we selected a standard unrelated set (also up to a 2^nd^ degree) which was used for all subsequent analyses (N=166,897). We further excluded individuals showing an excess of heterozygosity from the UK Biobank genotyping analysis and individuals of non-White British ancestry.

For the WMH analyses we used UKB field ID *25781* (N=38,347; N with WES=17,830; N unrelated=16,511). This study used the January 2020 release of UK Biobank imaging data on approximately 44,000 individuals. MRI was performed on two identical Siemens Skyra 3.0 T scanners (Siemens Medical Solutions, Germany), running VD13A SP4, with a standard Siemens 32-channel RF receiver head coil. Identical acquisition parameters and detailed quality control was used for all scans. Using the T2-FLAIR sequence, WMH volumes were generated by an image-processing pipeline developed and run on behalf of UKB and were available as part of the UK Biobank central analysis (https://biobank.ctsu.ox.ac.uk/crystal/crystal/docs/brain_mri.pdf) (Griffanti *et al*., 2016; Miller *et al*., 2016; Alfaro-Almagro *et al*., 2018). We excluded three extreme volume outliers (>6SD) and used log-transformed WMH volume to ensure normal distribution.

### Single Variant analysis

Single variant analysis of rare variants in the WES data was done using REGENIE (Mbatchou *et al*., 2020). REGENIE accounts for relatedness and subtle population stratification through a mixed-model approach. The mixed model parameters were estimated using 200,000 genotyped common variants. Saddle point approximation (SPA) regression was used in favor of Firth’s correction due to performance issues. We set a conservative exome-wide threshold of 5E-8 as significance cut-off in our analysis.

### Whole-exome burden test on white matter hyperintensities (WMH)

We determined the functional consequences of exome-wide variants using the Variant Effect Predictor (VEP) tool v101 (McLaren *et al*., 2016). For the whole-exome burden test on WMH analysis, we selected rare variants (MAF < 0.01) that are either predicted to be damaging by REVEL (Ioannidis *et al*., 2016) (REVEL score > 0.5) or predicted to exert a high-confidence loss-of-function effect using the LoFTEE (Karczewski *et al*., 2020) plugin in VEP.

For the domain specific analyses of *HTRA1*, we extracted all missense variants according to their position in the domain structure as defined by Uniprot identifier Q92743 (UniProt Consortium, 2019): signal peptide domain (Amino Acids [AA] 1-22), IGFBP domain (AA 33-100), Kazal-like domain (AA 100-157), Protease domain (AA 204-364) and PDZ domain (AA 365-467).

Whole-exome burden test were done using rvtests (Zhan *et al*., 2016) and by calculating multivariable burden tests using the CMC burden schema (Lee *et al*., 2014). Sex, age at imaging and 10 genomic principal components (PCs) were used as covariates in all analyses. For the primary logWMH analysis, we additionally corrected for vascular risk factors (SBP, LDL, BMI, Hba1C) in a sensitivity analysis. Results were deemed significant at an FDR level of 5% after FDR correction. To estimate effect sizes and standard errors, we used linear regression with minor allele carrier status as a dependent variable and age at imaging, sex and 10 genomic PCs as covariates.

To compare effect sizes to dichotomised risk factors for WMH, we extracted the following information from the UKB: hypertension status (yes/no), defined as a systolic blood pressure (SBP) >140 mmHg or a diastolic blood pressure (DBP) >90mmHg (UKB fields *4080* and *4079*) taking the mean of available measurements and correcting SBP and DBP for antihypertensive drug use (UKB field *2003*) as appropriate; diabetes mellitus (yes/no, UKB field *2443*); BMI (above mean vs. below mean; UKB field *21001*); smoking (ever smoked vs never smoked; UKB field *20116*). We further calculated a polygenic risk score (PRS) based on common genetic variants from the most recent WMH GWAS (Sargurupremraj *et al*., 2020). Specifically, we selected the 27 genome-wide significant lead SNPs and constructed a weighted allelic risk score for each individual in our dataset. Variants were weighted by their respective effect sizes in their association with WMH. We compared individuals above the mean vs below the mean, with a PRS >95% percentile vs PRS <95% percentile, and a PRS > 99.8% percentile vs PRS<99.8% percentile. The cut-off of 99.8% was chosen to achieve the same case number as for the HTRA1 protease domain variant carrier status. Thus, the standard error is identical.

### Recombinant protein expression

#### Expression vectors

The cDNA encoding human full-length HTRA1 (AA 1-480) or human full-length EGFL8 (AA 1-293, Origene) were cloned into a pcDNA4/TO/myc-His expression plasmid (Invitrogen). The cDNA encoding the N-terminal region of human LTBP1 (Latent-transforming growth factor beta-binding protein 1, AA 1-689) fused to a C-terminal V5-His tag was cloned into the pTT5 plasmid. Mutagenesis was conducted using the QuickChange Lightning Site-Directed Mutagenesis kit (Agilent Technologies).

#### Cell culture and transfection

Human embryonic kidney (HEK) 293E cells were grown in Dulbecco’s modified Eagle’s medium (DMEM) containing GlutaMAX, 10% (v/v) fetal calf serum (FCS), 100 U/ml penicillin and 100 μg/ml streptomycin (all from Invitrogen). Cells were transfected with Lipofectamine 2000 (Invitrogen) and maintained for 48 h in FCS-free DMEM before culture medium was collected and centrifuged for 10 min at 400 *g* to remove debris.

#### EGFL8 enrichment

Stably transfected HEK cells were selected using Zeocin (100 µg/ml, Invitrogen), then maintained in FCS-free DMEM for 4-5 days. Culture medium was collected and centrifuged at 1,000 *g* for 15 min, dialyzed at room temperature against 0.5 x phosphate-buffered saline (PBS) for 2 h and overnight at 4°C against 0.5 x PBS containing 200 mM NaCl. Medium was gently agitated for 1 h at room temperature in the presence of Talon resin (5 µl solution per ml medium, Clontech) and the resin subsequently transferred to a gravity flow column. After washes, EGFL8 was eluted with 100 mM EDTA in PBS and the eluate dialyzed overnight at 4°C against 1,000 volume of Tris 50 mM, NaCl 150 mM, pH 8.0.

#### HTRA1 purification

Human HTRA1 wild type (wt) lacking the N-terminal Mac domain (AA 158-480) was produced as described previously (Poepsel *et al*., 2015) with small modifications. N-termnially StrepII-tagged wt HTRA1 was affinity-purified with a strep-tactin resin material (IBA lifescieces) and subsequently subjected to size exclusion chromatography using a Superdex 200 preparation grade column (GE Healthcare) in 20 mM HEPES, 50 mM NaCl, pH 7.5.

### Protease activity assays

Medium from HEK cells overexpressing LTBP1 was treated with medium from cells overexpressing HTRA1 for 24 h at 37°C. LTBP1 proteolysis was evaluated by anti-V5 immunoblot. Relative protease activity was calculated as the ratio of cleaved tointact LTBP1 and was normalized to HTRA1 expression, assessed by anti-Myc immunoblot.

EGFL8 was treated with 100 or 500 nM purified wt HTRA1 for 24 h at 37°C. Where indicated, 5 µM of the HTRA1 inhibitor NVP-LBG976 (obtained from Novartis Pharmaceuticals) were added to the reaction (Grau *et al*., 2005). Substrate proteolysis was assessed by anti-Myc (EGFL8) immunoblot.

### SDS-PAGE and immunoblot

Proteins were separated by SDS-PAGE and transferred onto polyvinylidene difluoride membranes (Immobilon-P, Millipore). Membranes were blocked in Tris-buffered saline (TBS) containing 0.2% (v/v) Tween 20 and 4% (w/v) skim milk and probed with anti-Myc (Santa Cruz Biotechnology, #sc-40, 1:5,000), anti-V5 (Invitrogen, #R960-25, 1:10,000) or anti-HTRA1 (R&D Systems, #MAB2916, 1:5,000) primary antibodies prepared in TBS/Tween/milk. Detection was performed with Horseradish peroxidase (HRP)-coupled secondary antibodies (Dako), Immobilon Western kit reagents (Millipore) and the Fusion FX7 documentation system (Vilbert Lourmat). Signal intensity was quantified with ImageJ.

### Immunohistochemistry

Frozen human brain (frontal subcortex) from a 55-year-old individual with no known cerebrovascular disorder was provided by the Brain-Net Europe Biobank (Ludwig-Maximilians-University Munich) and analyzed according to the guidelines of the local ethical committees. For EGFL8 detection, ten μm tissue sections were thawed to room temperature (RT) and fixed for 20 min in 4% (w/v) paraformaldehyde then permeabilized and blocked in PBS added with 0.1% (v/v) Triton-X100 and 5% (w/v) bovine serum albumin (BSA) for 1 h at RT. Primary antibodies were diluted in PBS/Triton/BSA or PBS/BSA as follows: anti-EGFL8 Ab (Sigma-Aldrich, #HPA061173, 1:50), anti-collagen IV Ab (SouthernBiotech, #1340-01, 1:200), Cy3-coupled anti-α smooth muscle actin (SMA) Ab (Merck, #C6198, 1:100) and incubation was performed overnight at 4 °C. Following washing with PBS, sections were incubated with Alexa Fluor 488- or 647-coupled secondary antibodies (Abcam, 1:100) and 4’,6-Diamidin-2-phenylindol (DAPI, ThermoFisher Scientific) diluted in PBS for 1 h at RT. Tissue was washed, mounted with Fluoromount (Sigma-Aldrich), and images captured by confocal microscopy (LSM800, Zeiss).

### PheWAS

To explore the association of specific regions and variants in the full range of phenotypes encoded within the UK Biobank, we used the Phecode Map 1.2 to map UKB ICD10-codes to Phecodes (Wu *et al*., 2019). We used all ICD10 codes (main position, secondary position, death records) from the UKB. We excluded Phecodes with <100 cases and Phecodes that are male- or female-specific. Individuals were assigned a case status if >1 ICD10 code mapped to the respective Phecode. Individuals meeting the pre-specified exclusion criteria were removed from the analysis, otherwise the individual was assigned a control status. Given the known association between white matter hyperintensities and stroke, we further analyzed any stroke (AS), any ischemic stroke (AIS) and intracerebral hemorrhage (ICH) as defined by the algorithmically defined stroke outcomes (UKB fields *42006-42013*). Results were deemed significant at an FDR level of 5% after FDR correction. To approximate effect size in a logistic regression framework, we used minor allele carrier status as an independent variable and age at baseline, sex and 10 genomic PCs as covariates. We used Firth’s correction for unbalanced case/control ratios in our logistic regression analysis.

## RESULTS

We analyzed up to 17,830 individuals from the UK Biobank with both WES data and quantitative data on WMH volume available. Their demographic characteristics are presented in **Supp Table 1**.

### A predicted splice donor loss-of-function variant in GBE1 associates with WMH volume

In the single variant analysis using a mixed model approach and correcting for sex, age at imaging, and 10 principal components (PCs), we found one rare variant (MAF < 0.01) to be associated at genome-wide significance level (p<5E-8) with lower logWMH volume (beta=-0.74, se=0.13) in 17,830 individuals. This variant is a predicted splice donor loss-of-function (LoF) variant in *GBE1* (rs192044702, c.691+2T>C, MAF= 0.00131) encoding 1,4-alpha-glucan branching enzyme 1. We further found three single variants in *KLHL21* (rs891958177), *PARP2* (rs750147320), *and ABCC1* (rs748043154) to reach a suggestive threshold of p<5E-7 for association with logWMH volume (**Supp Table 2**).

### Whole-exome burden test reveals *HTRA1* as a risk gene for WMH burden

We next performed a whole-exome burden test using the Combined Multivariate and Collapsing (CMC) method implemented in the rvtests software. Analyses were corrected for sex, age at imaging, and 10 genomic PCs as covariates with logWMH volume as the dependent variable (N=16,511 unrelated individuals). The results were subsequently filtered to retain genes with >20 alternative alleles in the analyzed dataset. *HTRA1* was the only gene significantly associated at an FDR level of 5% (p=5.6E-6), based on 59 alternative *HTRA1* allele carriers (prevalence of ∼3.6/1000 in the UKB) at 18 genomic positions. Four additional genes including *GBE1, DCAKD, EGFL8* and *RGS12* showed suggestive association at an FDR level of 25% (**Table 1, Supp Table 3, Supp Fig 1**). To estimate the effects of carrier status on logWMH volume we performed linear regression analysis on log transformed WMH volume and minor allele carrier status, corrected for sex, age at imaging, and 10 genomic PCs. Carriers of rare variants in either *HTRA1, EGFL8* or *RGS12* showed increased WMH volumes compared to non-carriers (*HTRA1*: beta=0.36, se=0.09, p=0.0003; *EGFL8*: beta=0.22, se=0.06, p=9.6E-5; *RGS12*: beta=0.29, se=0.10, p= 0.003, respectively), whereas carriers of rare variants in *GBE1* (beta=-0.20, se=0.05, p=6.7E-5) and *DCAKD* (beta=-0.21, se=0.05, p=0.0002) showed lower WMH volumes. The results did not materially change when further correcting for vascular risk factors (SBP, LDL, Hba1C, BMI and smoking) (**Supp Table 4)** In an additional sensitivity analysis, we only considered LoF mutations in the exome-wide burden test. The associations of *GBE1, DCAKD* and *EGFL8* with WMH volume were of similar magnitude as in the combined missense and LoF analysis. (**Supp Table 5**) In contrast, the association of *HTRA1*, which was based on only 9 LoF variants, was greatly diminished (p=0.97), and therefore, *HTRA1* LoF variants were not considered in further analyses. There was no association with LoF alleles in *RGS12* (p=0.69).

**Table 1.**
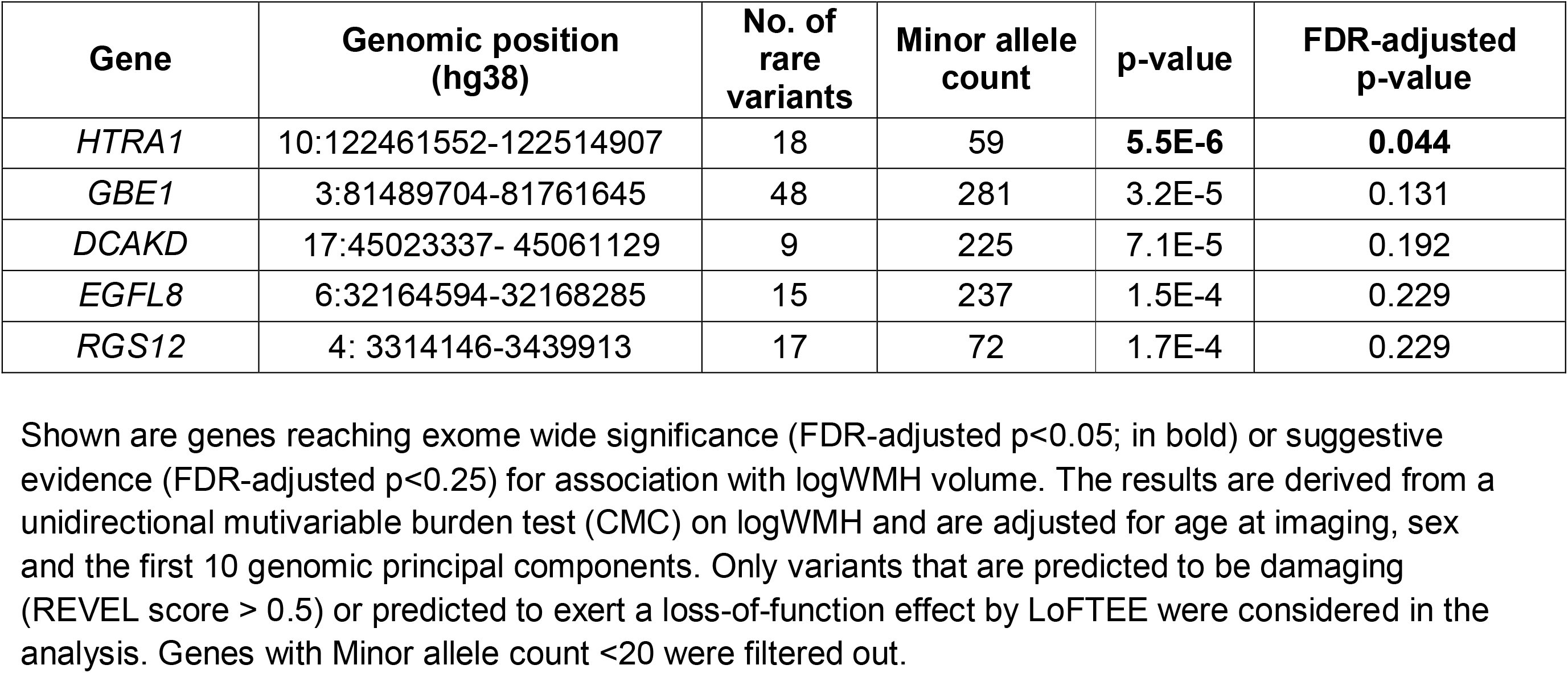
Whole-exome burden test on white matter hyperintensities.

| Gene | Genomic position (hg38) | No. of rare variants | Minor allele count | p-value | FDR-adjusted p-value |
| --- | --- | --- | --- | --- | --- |
| *HTRA1* | 10:122461552-122514907 | 18 | 59 | **5.5E-6** | **0.044** |
| *GBE1* | 3:81489704-81761645 | 48 | 281 | 3.2E-5 | 0.131 |
| *DCAKD* | 17:45023337- 45061129 | 9 | 225 | 7.1E-5 | 0.192 |
| *EGFL8* | 6:32164594-32168285 | 15 | 237 | 1.5E-4 | 0.229 |
| *RGS12* | 4: 3314146-3439913 | 17 | 72 | 1.7E-4 | 0.229 |

Shown are genes reaching exome wide significance (FDR-adjusted $p < 0.05$ ; in bold) or suggestive evidence (FDR-adjusted $p < 0.25$ ) for association with logWMH volume. The results are derived from a unidirectional multivariable burden test (CMC) on logWMH and are adjusted for age at imaging, sex and the first 10 genomic principal components. Only variants that are predicted to be damaging (REVEL score $> 0.5$ ) or predicted to exert a loss-of-function effect by LoFTEE were considered in the analysis. Genes with Minor allele count $< 20$ were filtered out.

### Domain-specific burden test of *HTRA1* shows specific association with the protease domain

Given the association between rare variants in *HTRA1* and WMH volume in the burden tests and the overall structure of the HTRA1 protease, which is organized into functionally distinct domains (**Fig 1**), we next repeated the analyses focusing on rare missense variants in individual domains. Specifically, we performed a separate CMC burden test for the signal peptide (AA 1-22, N=2 variants, prevalence 7.8/1000), IGFBP domain (AA 33-100, N=6 variants, prevalence 0.6/1000), Kazal-like domain (AA 100-157, N=4 variants, prevalence 0.9/1000), protease domain (AA 204-364, N=9 variants, prevalence 2.2/1000) and PDZ domain (AA 365-467, N=11 variants, prevalence 1.6/1000). Rare missense variants in the HTRA1 protease domain were significantly associated with altered WMH volumes (p=9.5E-8), whereas there was no signal for the other domains (**Fig 1** and **Table 2**) and the results remained stable after exclusion of one compound heterozygote individual (D320N / R403W). Baseline characteristics of protease domain variant carriers did not differ from those of non-carriers (**Supp Table 6**). We also calculated results for the combined protease and linker domain as the linker domain (AA 158-204; 2 variants) serves as an activator of protease function and found the association to be slightly attenuated (p=5.5E-7).

**Table 2.** *HTRA1* domain-specific burden test on white matter hyperintensities.

| Domain | Variants | MAC | beta | se | p-value |
| --- | --- | --- | --- | --- | --- |
| Signal peptide |  | 129 | -0.02 | 0.07 | 7.80E-01 |
|  | P4S | 2 | -0.21 | 0.61 | 7.34E-01 |
|  | A20V | 127 | -0.07 | 0.08 | 3.79E-01 |
| IGFBP domain |  | 11 | 0.18 | 0.23 | 4.30E-01 |
|  | A35S | 1 | 0.66 | 0.87 | 4.45E-01 |
|  | G36V | 3 | -0.92 | 0.50 | 6.54E-02 |
|  | P50A | 2 | -0.52 | 0.61 | 4.01E-01 |
|  | E51K | 1 | -1.53 | 0.87 | 7.77E-02 |
|  | G73D | 1 | -0.59 | 0.87 | 4.98E-01 |
|  | P82R | 3 | -0.42 | 0.50 | 4.06E-01 |
| Kazal domain |  | 15 | 0.12 | 0.22 | 5.20E-01 |
|  | R133G | 1 | 0.34 | 0.56 | 7.80E-01 |
|  | R138C | 1 | 0.23 | 0.66 | 4.50E-01 |
|  | Q151K | 12 | 0.02 | 0.25 | 9.34E-01 |
|  | Q157R | 1 | 0.32 | 0.87 | 7.17E-01 |
| Protease domain |  | 35 | 0.79 | 0.14 | **9.50E-08** |
|  | V221L | 1 | 1.87 | 0.87 | **3.10E-02** |
|  | V221M | 1 | 0.08 | 0.87 | 9.26E-01 |
|  | R227W | 20 | 0.73 | 0.19 | **1.71E-04** |
|  | K248T | 1 | 0.85 | 0.87 | 3.26E-01 |
|  | E272V | 3 | 0.03 | 0.50 | 9.59E-01 |
|  | P275L | 1 | 1.61 | 0.87 | 6.40E-02 |
|  | M314V | 1 | 0.48 | 0.87 | 5.83E-01 |
|  | D320N | 6 | 1.17 | 0.35 | **9.77E-04** |
|  | K362N | 1 | 1.82 | 0.87 | **3.60E-02** |
| PDZ domain |  | 26 | 0.06 | 0.17 | 4.30E-01 |
|  | A372V | 1 | 0.53 | 0.61 | 3.89E-01 |
|  | R403W | 3 | 1.01 | 0.50 | **4.51E-02** |
|  | V417I | 1 | 1.06 | 0.87 | 2.24E-01 |
|  | D420E | 4 | 0.26 | 0.43 | 5.49E-01 |
|  | S436G | 1 | -0.54 | 0.87 | 5.35E-01 |
|  | V442M | 1 | -0.96 | 0.87 | 2.71E-01 |
|  | A445T | 4 | 0.30 | 0.43 | 4.95E-01 |
|  | D450N | 2 | 0.01 | 0.61 | 9.83E-01 |
|  | V451I | 6 | -0.30 | 0.35 | 4.05E-01 |
|  | V461A | 1 | 0.36 | 0.87 | 6.75E-01 |
|  | R463C | 2 | -0.02 | 0.61 | 9.72E-01 |

The results are derived from a multivariable burden test (CMC, for domains) or from a Wald association statistic (for single variants). All analyses were adjusted for age at imaging, sex and the first 10 genomic principal components. All missense variants mapping to the respective domains were considered. Results are displayed for the specific domains (grey) or the single variants.

**Figure 1.**
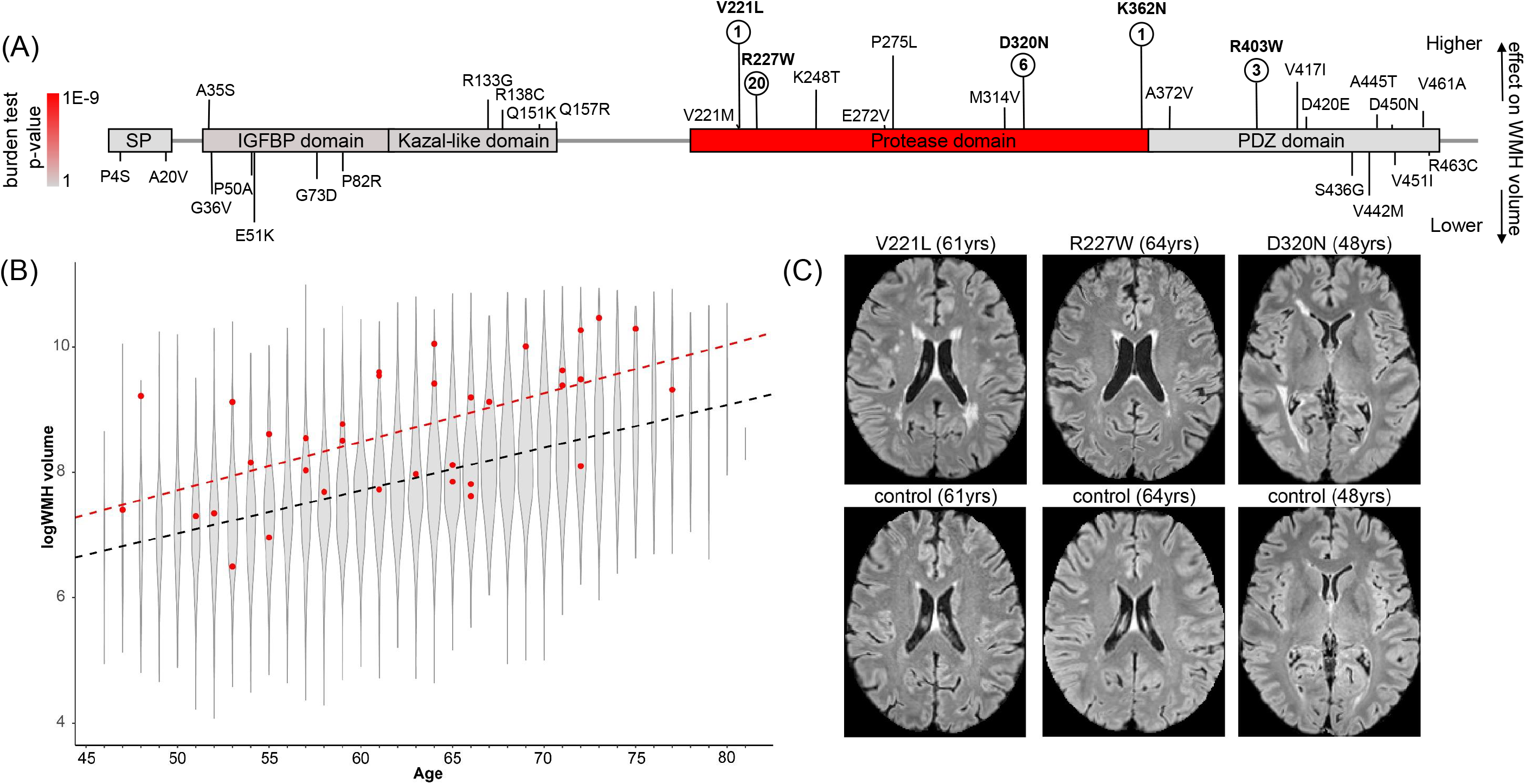
Overview of rare *HTRA1* variants and the association with WMH burden. (A) shown is the domain structure of the HTRA1 protein and the position of rare missense variants in the 16,511 UK Biobank participants included in the current analysis. Vertical lines reflect effect sizes (line length) and directionality of effect (increasing vs. decreasing) of the respective variant on WMH volume. Variants reaching statistical significance in single variant tests (p<0.05) are shown in bold with the number of minor allele carriers indicated below. (B) violin plot of age-stratified WMH burden in the UK Biobank. Carriers of a rare variant in the HTRA1 protease domain are depicted in red. The dashed lines represent fitted linear regression lines (WMH∼age) for all participants (black) and protease mutations carriers (red). (C) example brain MRI images of HTRA1 protease domain variant carriers and age-matched non-carriers.

To approximate the effects of carrier status on WMH volume, we performed linear regression analysis correcting for sex, age at imaging, and 10 genomic PCs. Carrier status of an alternative allele in the HTRA1 protease domain was associated with an increase in logWMH volume (beta=0.79, se= 0.14, p=9.4E-8; FDR <5%) whereas we found no effect of carrier status on WMH volume in other domains (**Table 2**). In an age-stratified analysis, carriers exhibited logWMH volumes comparable to non-carriers that were 11.4 years older (**Fig 1B**). When compared to other dichotomized risk factors for WMH volume, protease domain mutation carrier status showed a larger effect than hypertension (beta=0.26, se=0.02), diabetes (beta=0.25, se=0.04), BMI (beta=0.18, se=0.01) and smoking (beta=0.10, se=0.01). Also, comparison to individuals on a polygenic risk score (PRS) distribution based on 27 common genetic variants (Sargurupremraj *et al*., 2020) showed a larger effect size for carrier status of an alternative allele in the HTRA1 protease domain than for individuals with a PRS above vs below the 99.8% percentile of the PRS distribution (beta=0.44, se=0.14) (**Fig 2**).

**Figure 2.**
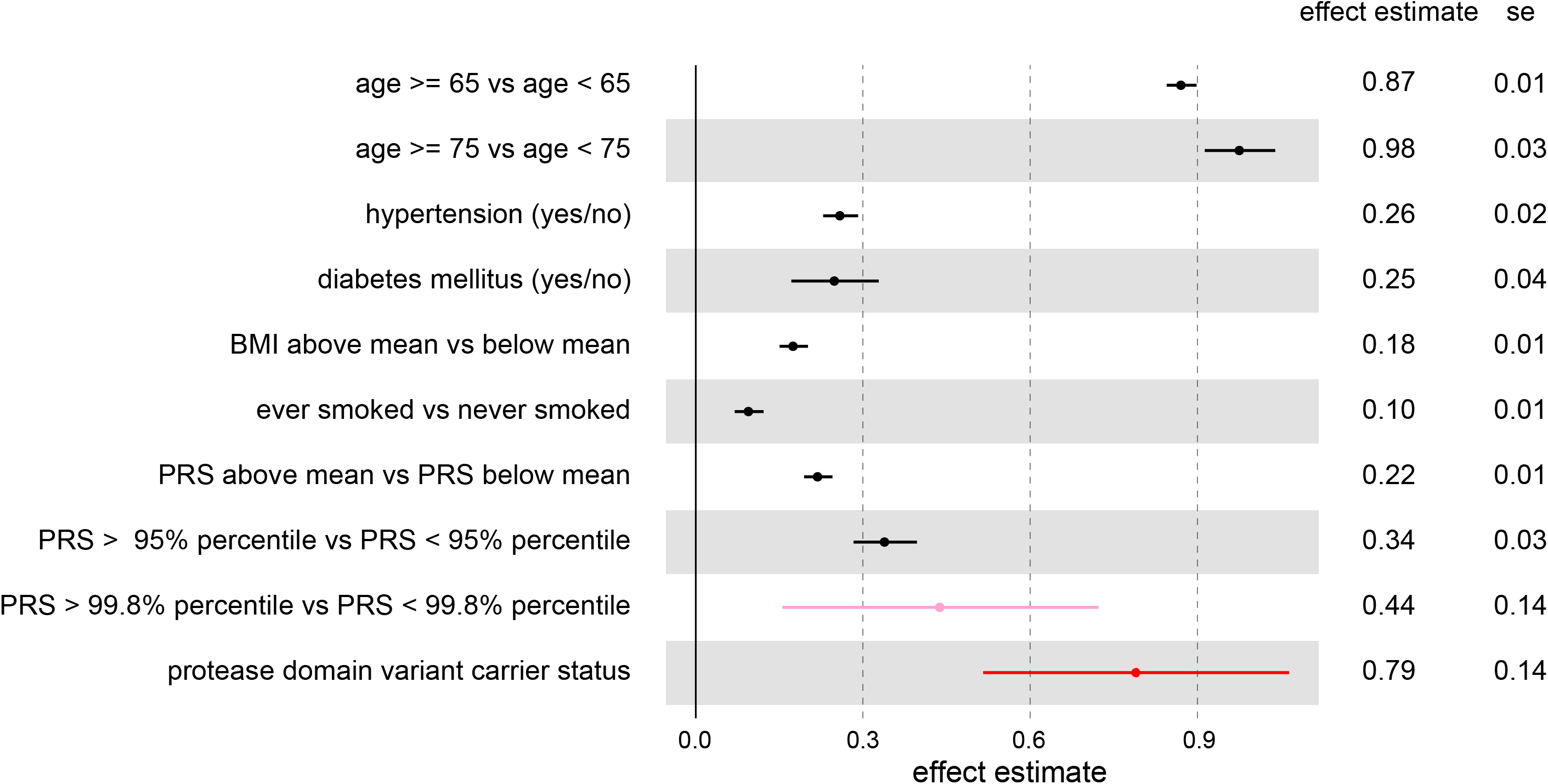
Effect size of protease domain variant carrier status on logWMH volume in comparison with established risk factors. Shown are the effect sizes and standard errors of dichotomized risk factors (age, hypertension, diabetes, smoking, BMI), a dichotomized polygenic risk score based on common genetic variants, and HTRA1 protease domain variant carrier status. Effect sizes and standard errors were derived for logWMH volume change using a linear regression model adjusting for age, sex and 10 genetic PCs.

### Mutations in the HTRA1 protease domain are associated with a loss of protease activity

To further elucidate the functional consequence of the identified *HTRA1* variants, we performed activity assays using human cell-derived recombinant proteins **(Fig 3)**. Specifically, we evaluated HTRA1 protease activity towards its known physiological substrate LTBP1. Wild type (wt) HTRA1 and an active site mutant (ASM) obtained by replacement of the catalytic Ser by an Ala (S328A) served as positive and negative controls, respectively. We investigated the enzymatic activity of 2 Kazal-like domain, 2 PDZ domain and 9 protease domain mutants. Three known CARASIL loss-of-function mutants were also included in the assays (Beaufort *et al*., 2014; Nozaki *et al*., 2016). We found the protease activity of Kazal-like and PDZ domain mutants to be comparable to that of wt HTRA1, in accord with the fact that these domains are dispensable for protease activity (Eigenbrot *et al*., 2012; Uemura *et al*., 2020). In contrast, 6 out of 9 protease domain mutants exhibited a marked reduction of protease activity (V221L, V221M, R227W, P275L, M314V, D320N).

**Figure 3.**
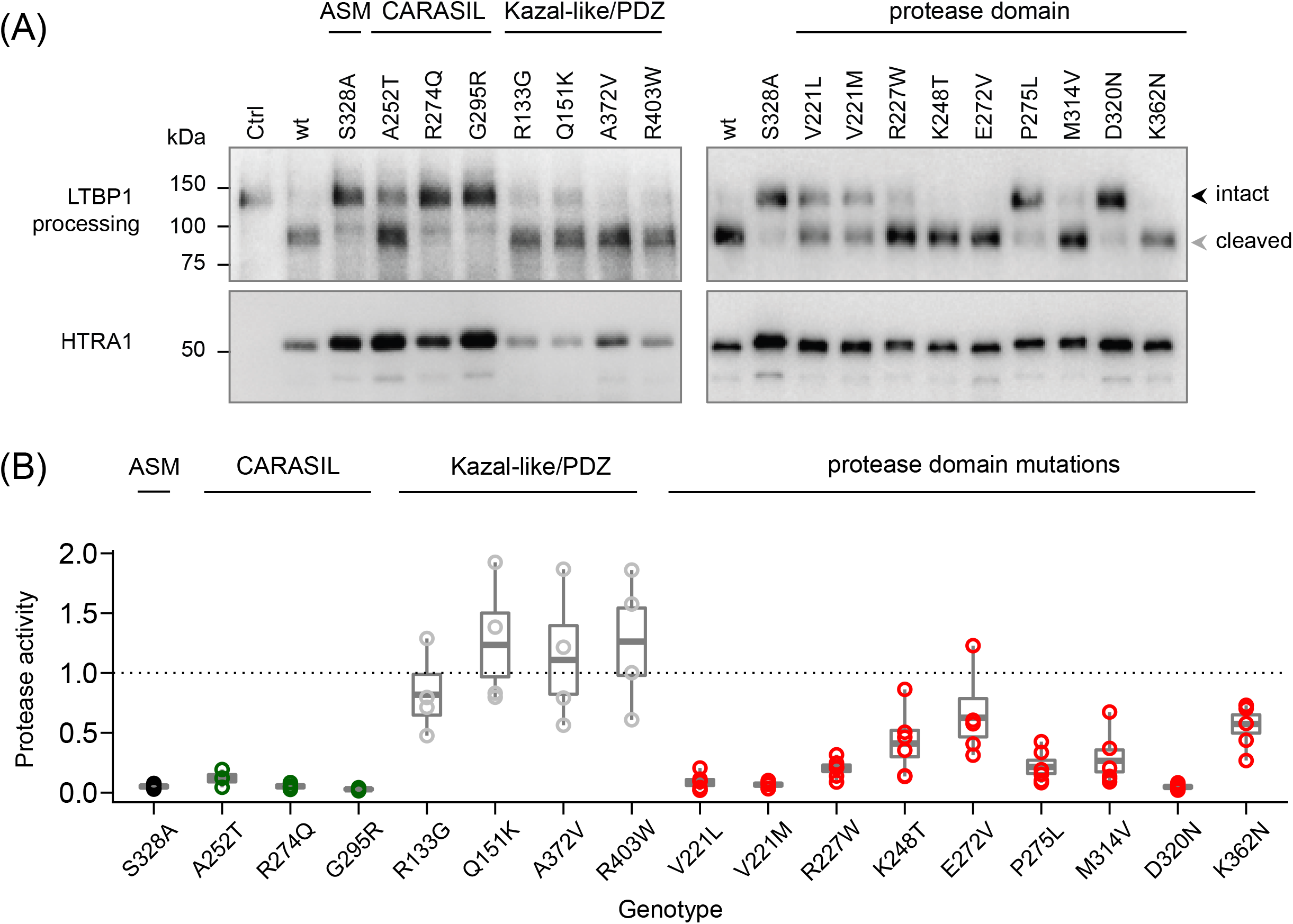
Functional consequences of rare *HTRA1* variants. (A) Culture medium from transfected cells was collected to assess HTRA1 expression (anti-Myc immunoblot) and protease activity towards LTBP1 (anti-V5 immunoblots). ASM: active site mutant. (B) Protease activity was determined as the ratio between cleaved (grey arrowhead) and intact (black arrowhead) LTBP1 and was normalized to HTRA1 levels. Data are presented as box-and-whisker plots (median +-standard error of mean; the activity of wt HTRA1 was set to 1). Data points represent independent measurements. At least three independent biological replicates were analyzed. ASM: active site mutant.

### EGFL8 is proteolytically processed by HTRA1 and expressed in the human brain vasculature

Given the prominence of variants in the HTRA1 protease domain in the association analyses and the result of the protease assays, we next performed a targeted analysis of genes encoding potential substrates of HTRA1. Potential substrates were identified from a recent proteomic study in Htra1 deficient mice (Zellner *et al*., 2018) that found 38 proteins to be enriched in the brain vasculature of *Htra1*^-/-^ compared to wild-type mice, among them several verified HTRA1 substrates including Efemp1, Vtn, and Ltbp1 (Lin *et al*., 2018; Zellner *et al*., 2018) (**Supp Table 7**). Notably, one of the most strongly enriched proteins in that dataset was Egfl8. Focusing on the corresponding genes encoding these 38 candidate substrates in our burden test, *EGFL8* was the only gene that reached experiment-wide significance at an FDR level of 5% (p=1.54E-4).

We next set out to experimentally determine whether the human EGFL8 protein is a substrate of HTRA1. Treatment of EGFL8 with HTRA1 resulted in a dose-dependent degradation of EGFL8, which was prevented in the presence of a selective HTRA1 inhibitor **(Fig 4A)**. These observations thus identify EGFL8 as a novel HTRA1 substrate. We further examined the cerebral expression pattern of EGFL8 by immunohistochemistry. In the human cortex, EGFL8 was predominantly localized to the alpha smooth muscle actin positive vasculature, thus corresponding to arteries and arterioles (**Fig 4B**).

**Figure 4.**
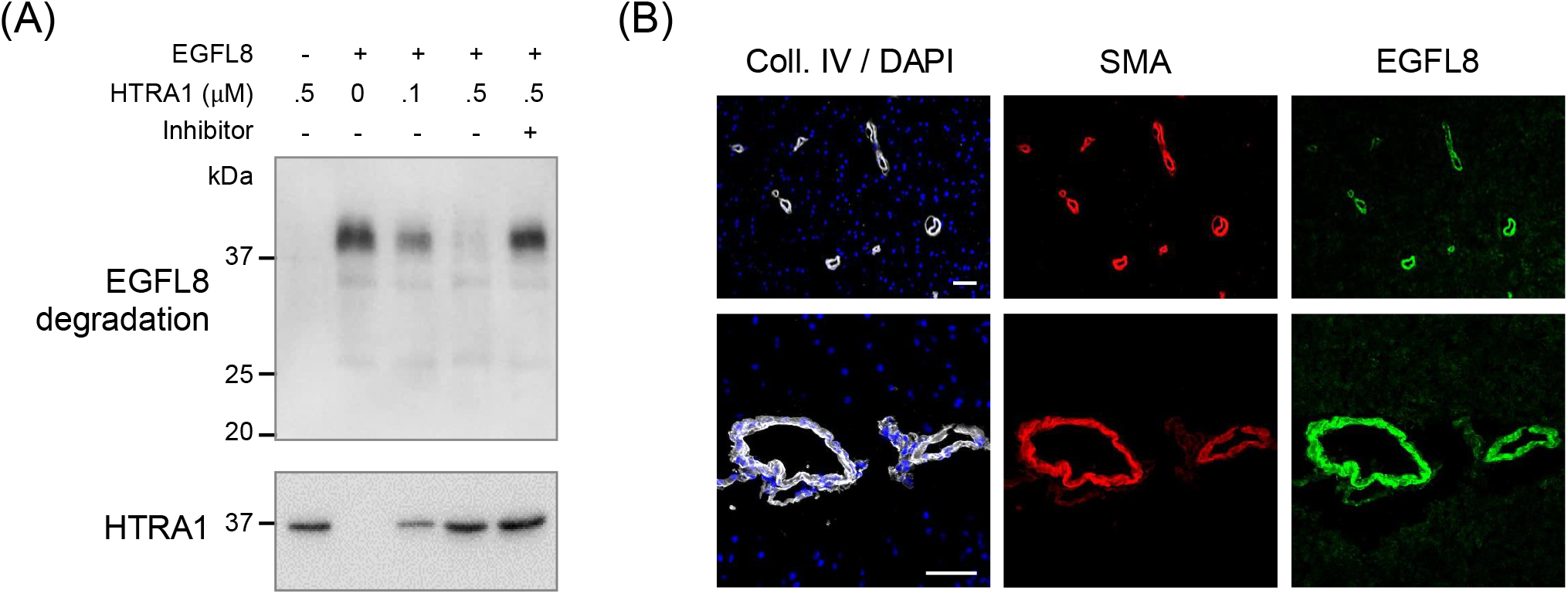
EGFL8 is a direct substrate of HTRA1 and expressed in the human brain vasculature. (A) EGFL8 was exposed to increasing concentrations of purified HTRA1 in the absence or presence of an HTRA1 inhibitor. EGFL8 and HTRA1 were detected by anti-Myc and anti-HTRA1 immunoblots, respectively. (B) Immunohistochemical detection of EGFL8 in human brain sections. The vascular basement membrane component collagen IV (Coll. IV) and the smooth muscle cell marker alpha smooth muscle actin (SMA) were detected as controls. Shown are representative images. Scale bar: 50µm.

### Rare variants in the HTRA1 protease domain associate with multiple neurological and non-neurological phenotypes

*HTRA1* mutation carriers exhibit a wide range of cerebral and extracerebral phenotypes (Rannikmae *et al*., 2020). To comprehensively assess disease outcomes associated with rare variants in *HTRA1* or *EGFL8*, we performed a rare-variant burden PheWAS on standardized Phecodes in the UK Biobank. Specifically, we matched patients’ ICD10 codes (both primary, secondary and death-record based) to PheCodes. After excluding Phecodes that were sex-specific or had <100 cases in the analysis, 741 different Phecodes remained that were analyzed in burden tests. We further performed analyses for any stroke (AS), any ischemic stroke (AIS) and intracerebral hemorrhage (ICH) as defined by the algorithmically defined stroke outcomes (UKB fields *42006-42013*) and for parental history of stroke and family history of dementia (UKB field *20107* and *20110*).

Considering all identified damaging and LoF variants in *HTRA1* (**Supp Tables 8** and **9**, we found conductive hearing loss (Phecode 389.2) to be associated at an FDR level of 5% (**Supp Table 10, Supp Fig 2**) with minor allele carriers having a higher risk in Firth logistic regression (OR: 5.37, CI95 [2.39-9.49]). Focusing on missense variants in the protease domain we identified 12 Phecodes at an FDR level of 5% (**Fig 5, Table 3 and Supp Table 11**) including migraine with aura (Phecode 340.1), encephalitis (Phecode 323), aneurysms and dissection of heart (Phecode 411.41), and heart valve replaced (Phecode 395.6). Notably, for all significant Phecodes minor allele carrier status was associated with higher risk of the respective phenotype. For *EGFL8*, we identified two Phecodes at an FDR level of 5%: 578.8 (Hemorrhage of rectum and anus) and 696.41 (Psoriasis vulgaris), the latter confirming prior work in the UKB (Emdin *et al*., 2018) (**Supp Table 12, Supp Fig 3**).

**Table 3.** *HTRA1* protease domain PheWAS.

| Phecode | Phenotype | P-value | FDR-Adjusted P-value | Odds Ratio | 95% Confidence Interval |
| --- | --- | --- | --- | --- | --- |
| 389.2 | Conductive hearing loss | 3.60E-06 | 0.0025 | 10.75 | 2.99-26.90 |
| 571.6 | Primary biliary cirrhosis | 3.37E-05 | 0.012 | 13.37 | 2.77-38.77 |
| 411.41 | Aneurysm and dissection of heart | 6.34E-05 | 0.015 | 6.68 | 2.22-15.20 |
| 340.1 | Migraine with aura | 8.31E-05 | 0.015 | 12.24 | 2.54-35.25 |
| 395.6 | Heart valve replaced | 0.00011 | 0.016 | 4.70 | 1.92-9.49 |
| 323 | Encephalitis | 0.00016 | 0.019 | 11.43 | 2.38-32.92 |
| 291.8 | Alteration of consciousness | 0.00018 | 0.019 | 7.68 | 2.14-19.13 |
| 324 | Other CNS infection and poliomyelitis | 0.00027 | 0.024 | 10.80 | 2.24-31.13 |
| 575.1 | Cholangitis | 0.00042 | 0.032 | 7.08 | 1.97-17.68 |
| 613.1 | Inflammatory disease of breast | 0.00044 | 0.032 | 7.04 | 1.96-17.64 |
| 857 | Mechanical complication of unspecified genitourinary device, implant, graft | 0.00055 | 0.036 | 4.62 | 1.74-9.78 |
| 465 | Acute upper respiratory infections of multiple or unspecified sites | 0.00067 | 0.040 | 4.53 | 1.70-9.58 |

Results from a PheWAS analysis using Phecodes as the phenotypes of interest. Shown is the association of variants in the protease domain of *HTRA1* with the individual Phecodes in a CMC burden test. Odds Ratios were calculated using Firth's regression and rare allele carrier status as an independent variable. FDR: false discovery rate.

**Figure 5.**
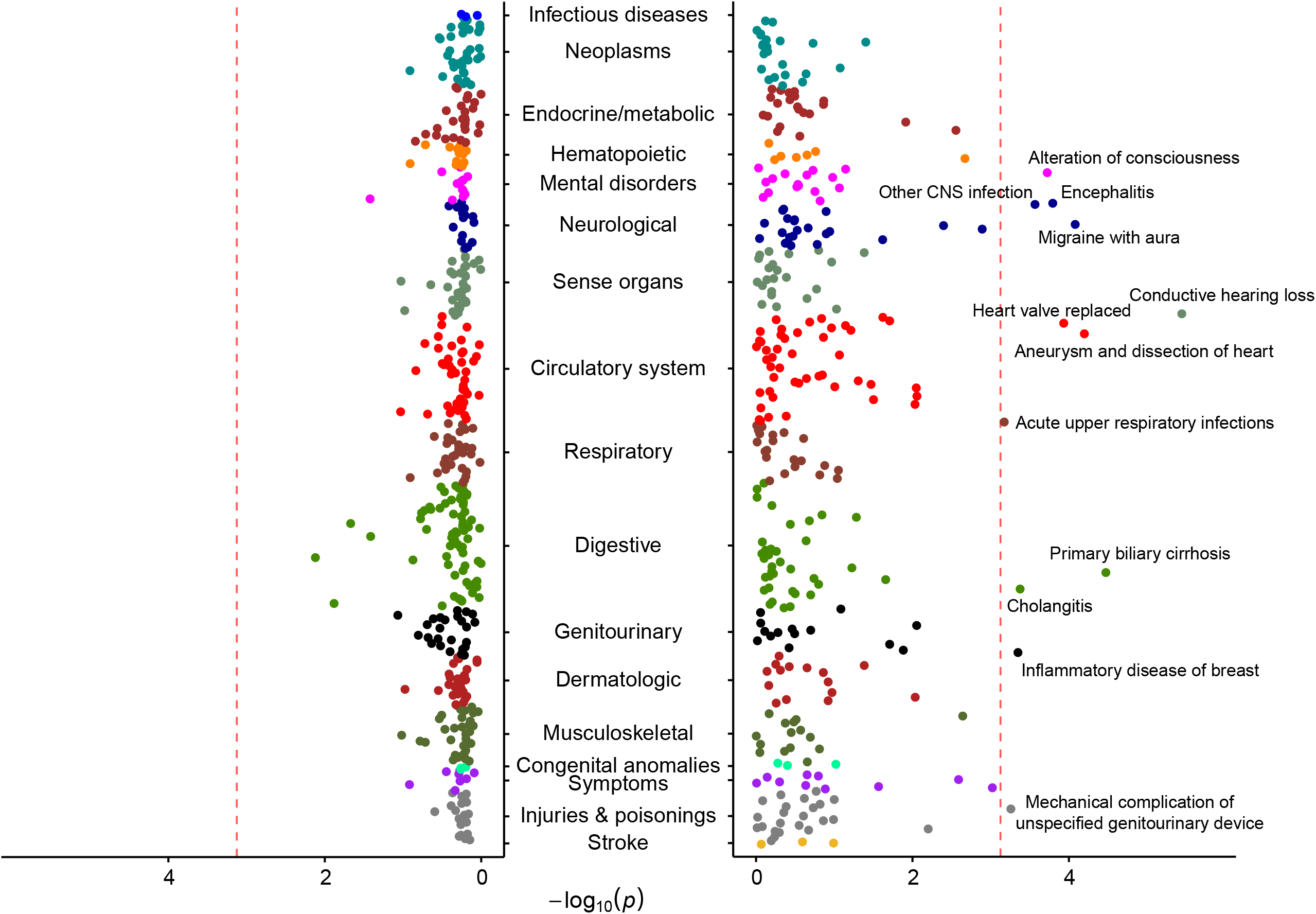
Rare variants in the HTRA1 protease domain associate with multiple neurological and non-neurological phenotypes. Phenome-wide association study (PheWAS) Manhattan plot for carriers of rare variants in the HTRA1 protease domain. Shown are all standardized Phecodes along with stroke-related phenotypes. For phenotypes to the right, HTRA1 protease domain variant carriers show higher risk, for phenotypes to the left, carriers are at lower risk. The red dashed line represents the FDR 5% cut-off.

## DISCUSSION

Using whole-exome sequencing data from the UK Biobank, we show that rare variants in *HTRA1* are associated with a higher burden of radiological white matter hyperintensities in the general population. We further show that the association with WMH burden is largely restricted to variants affecting the HTRA1 protease domain and provide experimental evidence that variants giving the strongest association signal result in a loss of proteolytic activity of HTRA1. Our PheWAS results reveal that heterozygous variants in the HTRA1 protease domain, which are present at a frequency of 1 in 450 in the UKB population, are associated with a wide range of phenotypes including migraine with aura. We further demonstrate that the gene product of *EGFL8*, which shows suggestive association with WMH burden in the UKB, is a direct substrate of HTRA1 and is expressed in the brain vasculature. Collectively, these findings highlight an important role of the HTRA1 protease in maintaining white matter integrity and provide insights in disease mechanisms.

HTRA1 has been implicated in familial small vessel disease (SVD). Biallelic mutations in *HTRA1* cause cerebral autosomal recessive arteriopathy with subcortical infarcts and leukoencephalopathy (CARASIL), an early-onset condition that is characterized by stroke, dementia, spondylosis deformans, and alopecia (Hara *et al*., 2009; Beaufort *et al*., 2014). In contrast, heterozygous mutations in *HTRA1* cause an autosomal dominantly inherited SVD that displays a more restricted phenotype of stroke, cognitive decline, and gait disturbance and manifests at a later age of onset (Verdura *et al*., 2015). Our results extend these findings by showing that rare variants in *HTRA1*, and more specifically variants in the HTRA1 protease domain, are associated with WMH burden on a population-wide level. Notably, we found no association of such variants with stroke, dementia or a family history of stroke or dementia in the UKB. Instead, we found an association with both neurological and non-neurological phenotypes that have not previously been associated with *HTRA1* mutations.

The domain-specific analyses for HTRA1 in combination with the protease assays suggest that the mechanism underlying the observed association between rare variants in *HTRA1* and WMH burden is a loss of proteolytic function of the HTRA1 protease. This would be in line with the proposed mechanisms for mutations implicated in *HTRA1*-related familial SVD (Beaufort *et al*., 2014; Verdura *et al*., 2015). However, our experiments focused on a set of variants identified in the UKB and were limited to biochemical assays using recombinant proteins. Other putative loss-of-function mechanisms related to e.g., gene expression, mRNA or protein stability (Hara *et al*., 2009; Fasano *et al*., 2020) were not evaluated. Also, we did not go further in examining possible dominant negative effects, as recently reported for a subset of *HTRA1* variants linked to hereditary SVD (Nozaki *et al*., 2016; Uemura *et al*., 2020). Of interest, one of the variants (D320N) detected here has previously been reported in an Asian CARASIL patient (Xie and Zhang, 2018) who was compound heterozygous for D320N. In contrast, R227W, which had an allele frequency of 0.04% in the UKB population with available WES data (0.06% in those with imaging), was associated with WMH burden and has so far not been linked to neurological disease. More work is needed to delineate genotype-phenotype correlations at this locus.

While *DCAKD* and *EGFL8* did not reach exome-wide significance for association with WMH burden, we consider these genes to exhibit strong evidence for a causal involvement in WMH: Recent GWAS found common variants at *DCAKD* to reach genome-wide significance for association with WMH while leaving the responsible gene unidentified (Persyn *et al*., 2020; Sargurupremraj *et al*., 2020). The current study pinpoints *DCAKD* as a likely causal gene in this region and further demonstrates a role of rare variants in *DCAKD*. A causal involvement of *EGFL8* is supported by the following observations. First, we found the gene product of *EGFL8* to be efficiently cleaved by HTRA1 in our protease assays, thus demonstrating a functional interaction between EGFL8 and a protease with an established role in SVD and cerebral WMH. Notably, common variants in *EFEMP1* encoding another substrate of HTRA1 (Lin *et al*., 2018; Armstrong *et al*., 2020; Persyn *et al*., 2020; Sargurupremraj *et al*., 2020), have consistently been associated with WMH in recent GWAS (Lin *et al*., 2018; Armstrong *et al*., 2020; Persyn *et al*., 2020; Sargurupremraj *et al*., 2020). Second, we found human EGFL8 to be prominently expressed in the brain vasculature, which would be in line with the role of vascular mechanisms in WMH. The biological function of *EGFL8*, encoding ‘EGF Like Domain Multiple 8’, remains largely unexplored. A recent study (Weiss *et al*., 2021) found EGFL8 to act as neuritogen and to rewire cellular signaling by activating kinases involved in neurogenesis. Notably, *EGFL7*, a close paralogue of *EGFL8*, which is likewise expressed in brain vessels, has been implicated in various vascular functions including angiogenesis and elastogenesis (Parker *et al*., 2004; Lelievre *et al*., 2008; Nichol and Stuhlmann, 2012). *DCAKD*, encoding ‘Dephospho-CoA Kinase Domain Containing protein’, is ubiquitously expressed in brain, has a putative role in neurodevelopment, and has previously been shown to be implicated in psychiatric disease (Bipolar Disorder Schizophrenia Working Group of the Psychiatric Genomics Consortium, 2018; Barbu *et al*., 2020). Moreover, studies on expression quantitative loci (eQTL) have linked expression levels of *DCAKD* to white matter microstructure and WMH volume (Barbu *et al*., 2020; Persyn *et al*., 2020).

Among the phenotypes shown here to be associated with rare variants in the HTRA1 protease domain is migraine with aura thus pointing to a potential mechanistic link between WMH and migraine with aura. Previous population-based studies found a higher frequency of radiological WMH in migraineurs (Kruit *et al*., 2004; Kurth *et al*., 2011; Palm-Meinders *et al*., 2012; Hamedani *et al*., 2013). However, this was not consistently observed in other studies (Monteith *et al*., 2014; Gaist *et al*., 2016) and there was no clear pattern in terms of a preferential association with migraine with aura compared to migraine without aura (Kruit *et al*., 2004; Kurth *et al*., 2011; Palm-Meinders *et al*., 2012; Hamedani *et al*., 2013). Of further interest, common variant association studies reveal that several of the risk loci for migraine overlap with known risk loci for WMH including *COL4A1, COL4A2, NBEAL1-CARF*, and *ADAMTSL4* (Persyn et al., 2020; Sargurupremraj et al., 2020; Hautakangas et al., 2021). The association between rare variants in *HTRA1* and migraine with aura observed here further adds to recent GWAS that found common variants at *HTRA1* to be associated with migraine (Gormley *et al*., 2016; Hautakangas *et al*., 2021). Studies in additional datasets are needed to better delineate the relationship between rare variants in *HTRA1* and migraine and a potential mechanistic link with WMH.

The setting of the UKB enabled us to provide an estimate of the frequency of rare variants and their impact on WMH on a population level. Altogether, we found rare variants in the HTRA1 protease domain to occur at a frequency of about 1 in 400. As expected, some variants, such as R227W, were found to be more prevalent than others and effect sizes varied between variants although these results need to be interpreted with caution given the low number of risk allele carriers. Contrasting with the current gene-burden tests that considered rare variants in *HTRA1*, common variants at *HTRA1* did not reach genome-wide significance for association with WMH in two recent GWAS that mostly included individuals from the general population (Persyn *et al*., 2020; Sargurupremraj *et al*., 2020). However, a recent candidate gene association study in population-based subjects with extreme WMH volumes found a common intronic variant in *HTRA1* (rs2293871; allele frequency: 19%) to be associated with WMH volume (Mishra *et al*., 2019). Also, *HTRA1* reached genome-wide significance in a multi-trait analysis that combined GWAS data on WMH and lacunar stroke (Traylor *et al*., 2021). Overall, these findings emphasize the relevance of *HTRA1* for vascular brain injury beyond rare hereditary arteriopathies.

Our single variant analysis revealed one variant that reached exome-wide significance and three additional variants that reached suggestive evidence for association with WMH. rs192044702 in *GBE1* has previously been reported in compound heterozygous individuals with glycogen storage disease IV (Fernandez *et al*., 2010; Ravenscroft *et al*., 2013), which is allelic to adult polyglucosan body disease (APBD, OMIM #263570) (Mochel *et al*., 2012). The majority of patients with APBD have WMH on MRI (Mochel *et al*., 2012; Lopez Chiriboga, 2017), which seems at odds with our observation of a protective effect of rs192044702 on WMH burden. However, carriers of a single APBD mutation are generally asymptomatic and the effects by which variants in GBE1 might influence white matter integrity in one or the other direction remain poorly understood.

Our study has limitations. First, while the analysis of a quantitative trait offers higher statistical power than a binary phenotype our study still had limited power to detect small to moderate effect sizes. Also, the low number of alternative allele carriers may have resulted in inaccurate effect estimates with wide confidence intervals and potentially also false-positive findings. Second, as with any WES approach, there remains a possibility of errors in calling very rare variants. However, this would be expected to bias the results from gene burden tests towards the null hypothesis whereas we found a high consistency in the directionality of effects for associated variants in *HTRA1*. Also, despite rigorous QC of WES data in the UKB (Szustakowski *et al*., 2020), we cannot exclude the possibility of mis-called ultra-rare variants. Third, the results from our PheWAS might be more favorably powered for phenotypes with an early age-of onset. Given the relatively young age of UKB participants at study inclusion (mean age 56.8yrs) and the low number of participants with long-term follow up (median 8.98yrs) associations with late-onset phenotypes such as stroke and dementia might have been missed. As a possible indication we found rare variants in the HTRA1 protease domain to only reach nominal significance for Phecodes representing “transient cerebral ischemia” (Phecode 433.21) and “occlusion of cerebral arteries” (Phecode 433.31). Fourth, the level of phenotyping for stroke in the UKB precluded analyses restricted to lacunar stroke. Fifth, our PheWAS analysis was limited to hospital episode stay and death record ICD10 codes mapped to Phecodes. Several symptoms and phenotypes of interest (e.g. alopecia) are not sufficiently reflected by these codes. Sixth, we did not assess longitudinal changes in WMH burden as the number of individuals with available follow-up data on WMH in the UKB is still very low (approx. 1,500 individuals). Finally, our results cannot be extrapolated to other ethnicities and populations, which differ in terms of genetic architecture, vascular risk factors, and environmental factors (Brickman *et al*., 2008; Zahodne *et al*., 2015; Sargurupremraj *et al*., 2020).

## Data Availability

N/A

