## Supplementary Information for "Whole-exome sequencing in 16,511 individuals reveals a role of the HTRA1 protease and its substrate EGFL8 in brain white matter hyperintensities"

**SUPPLEMENTARY MATERIAL**

**Supplementary Figures**

**Supplementary Figure 1. Manhattan plot of the CMC burden test on logWMH volume.** Plotted are the genomic position (x-axis) and the -log10 p-value (y-axis) for each gene analyses. The red line indicates a false discovery rate (FDR) < 5%, the blue line indicates a FDR < 25%.

**Supplementary Figure 2. PheWAS plot for *HTRA1* considering all identified damaging and LoF variants.** Plotted are, for all 741 standardized Phecodes, the CMC association burden p-value (x-axis). The y-axis represents the grouping of the Phecodes. Data points to the left show decreased risk with variant carrier status, while data points to the right show increased risk for the phenotype of interest with variant carrier status.

**Supplementary Figure 3. PheWAS plot for *HTRA1* protease domain variants.** Plotted are, for all 741 standardized Phecodes, the CMC association burden p-value (x-axis). The y-axis represents the grouping of the Phecodes. Data points to the left show decreased risk with variant carrier status, while data points to the right show increased risk for the phenotype of interest with variant carrier status.

**Supplementary Tables**

Please see enclosed XLS file for Supplementary Tables 1-12
