## Supplementary figures and images for "Whole-exome sequencing in 16,511 individuals reveals a role of the HTRA1 protease and its substrate EGFL8 in brain white matter hyperintensities"

### Supp Fig 1

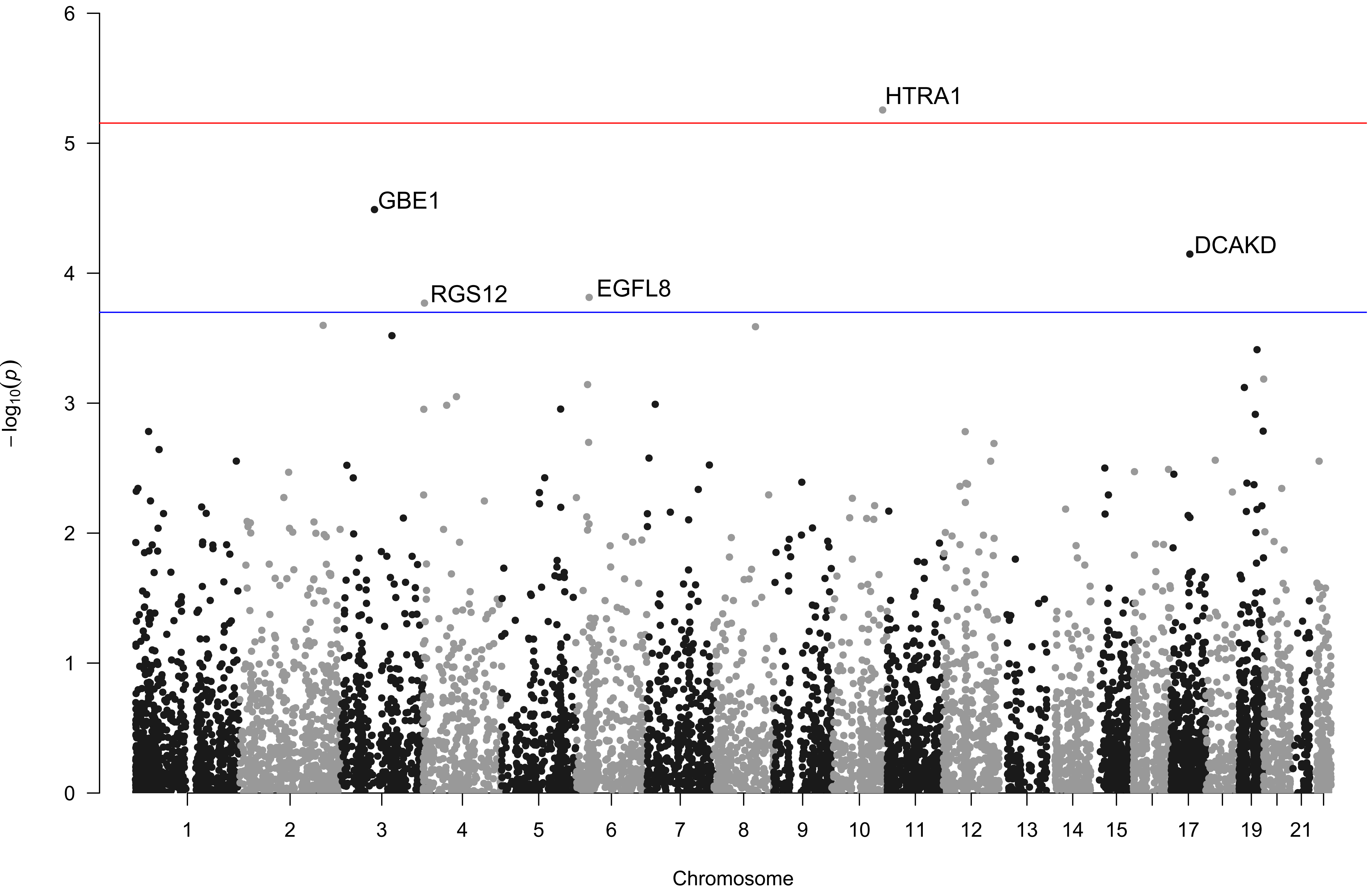

### Supp Fig 2

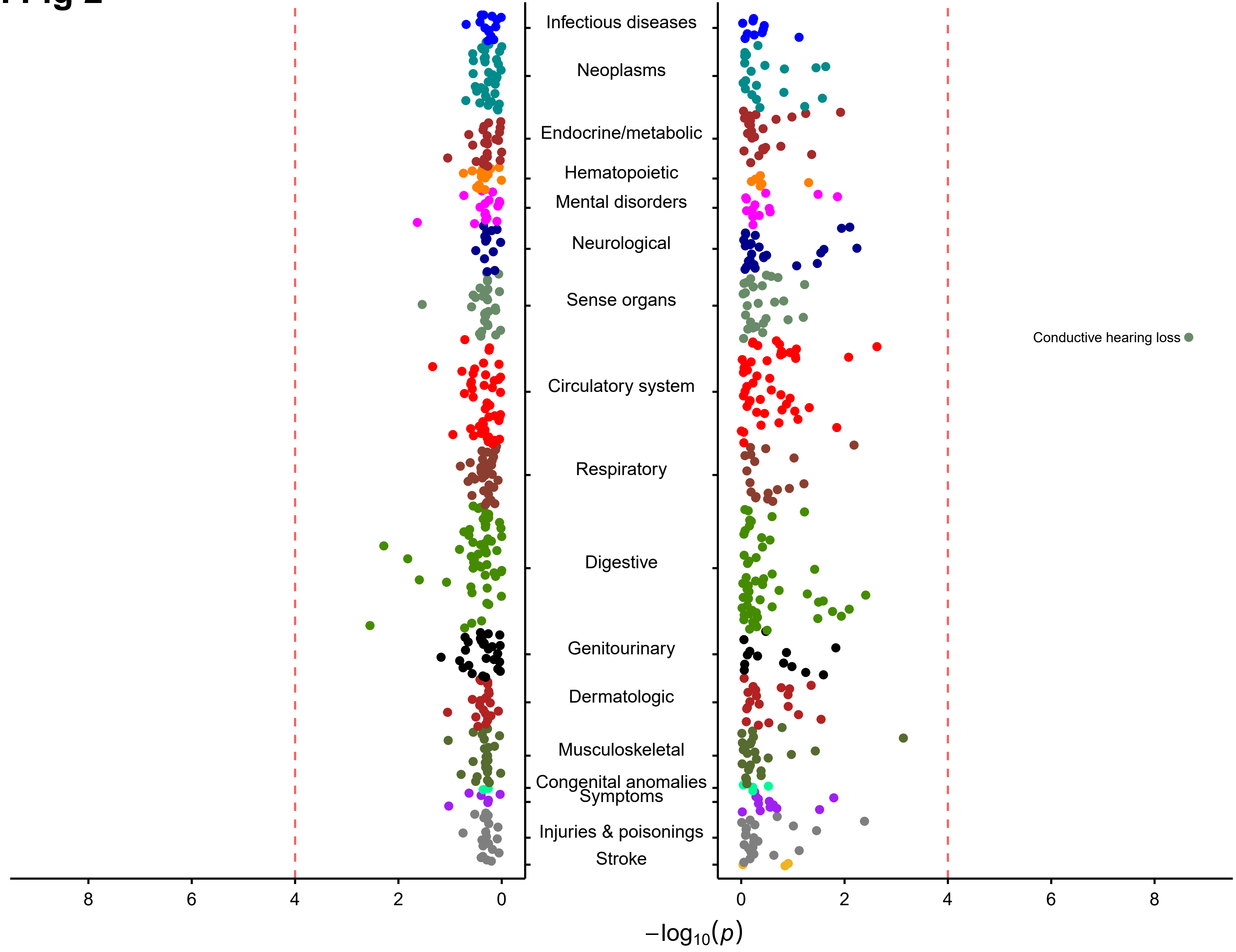

### Supp Fig 3

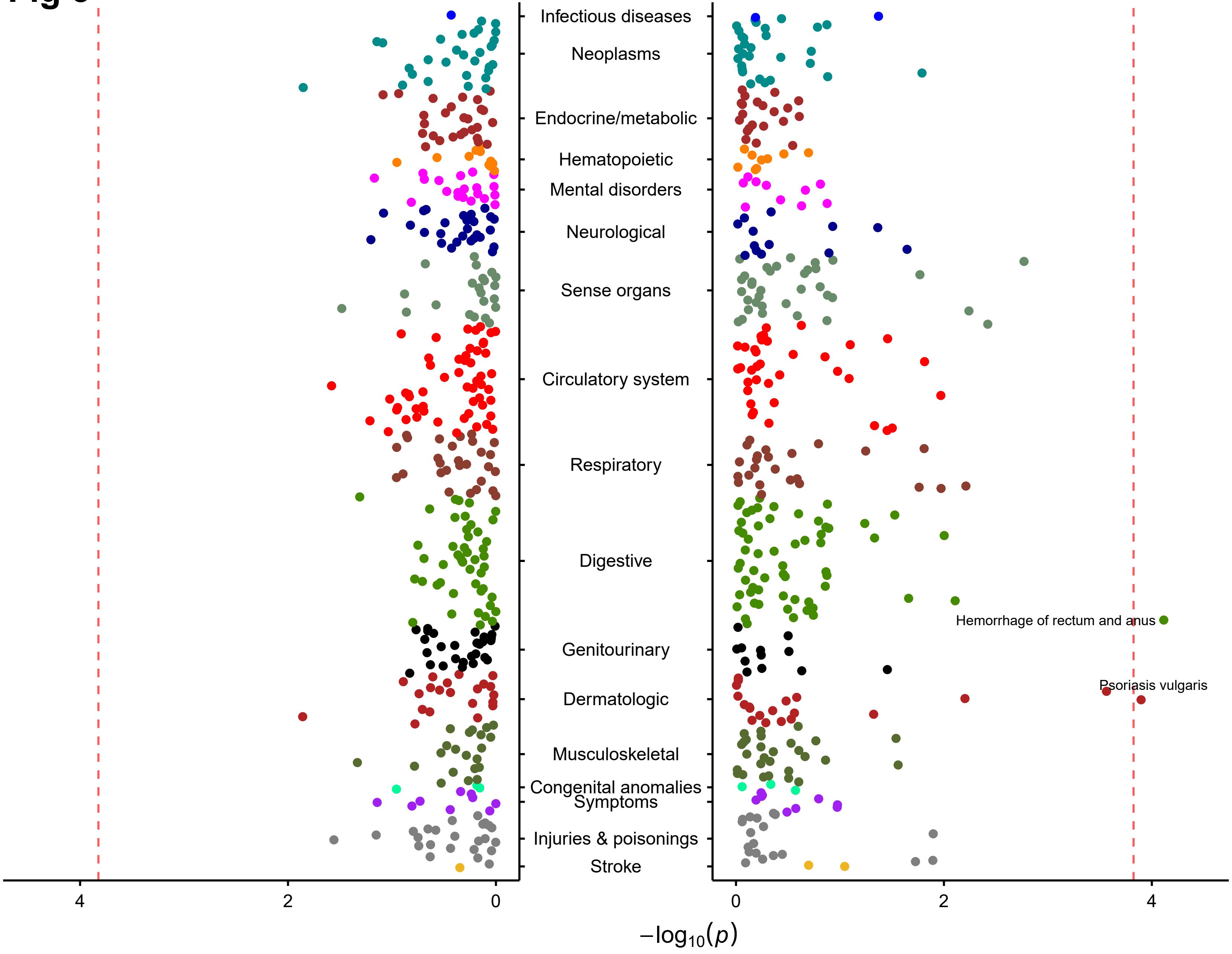
